# EyeG2P: an automated variant filtering approach improves efficiency of diagnostic genomic testing for inherited ophthalmic disorders

**DOI:** 10.1101/2021.07.23.21261017

**Authors:** Eva Lenassi, Ana Carvalho, Anja Thormann, Tracy Fletcher, Claire Hardcastle, Sarah E Hunt, Panagiotis I Sergouniotis, Michel Michaelides, Andrew R Webster, F Cunningham, Simon Ramsden, David R FitzPatrick, Graeme CM Black, Jamie M Ellingford

**Affiliations:** Division of Evolution and Genomic Sciences, School of Biological Sciences, Faculty of Biology, Medicine and Health, University of Manchester, Manchester, United Kingdom; Manchester Centre for Genomic Medicine, St Mary’s Hospital, Manchester University NHS Foundation Trust, Manchester, United Kingdom; Manchester Royal Eye Hospital, Manchester University NHS Foundation Trust, Manchester, United Kingdom; MRC Human Genetics Unit, MRC Institute of Genetics and Cancer, University of Edinburgh, Edinburgh, UK; Medical Genetic Unit, Pediatric Hospital, Coimbra Hospital and Universitary Centre (CHUC), Coimbra, Portugal; European Molecular Biology Laboratory, European Bioinformatics Institute, Wellcome Genome Campus, Hinxton, CB20 1SD, Cambridge, UK; UCL Institute of Ophthalmology, University College London, London, UK; Department of Ophthalmology, Moorfields Eye Hospital NHS Foundation Trust, London, UK

## Abstract

**Purpose:** The widespread adoption of genomic testing for individuals with ophthalmic disorders has increased demand on diagnostic genomic services for these conditions. Moreover, the clinical utility of a molecular diagnosis for individuals with inherited ophthalmic disorders is increasingly placing pressure on the speed and accuracy of genomic testing.

**Methods:** We created EyeG2P, a publically available resource to assist diagnostic filtering of genomic datasets for ophthalmic conditions, utilising the Ensembl Variant Effect Predictor. We assessed the sensitivity of EyeG2P for 1234 individuals with a broad range of conditions, who had previously received a confirmed molecular diagnosis through routine genomic diagnostic approaches. For a prospective cohort of 83 individuals, we also assessed the precision of EyeG2P in comparision to routine genomic diagnostic approaches.

**Results:** We observed that EyeG2P had a 99.5% sensitivity for genomic variants previously identified as a molecular diagnosis for 1234 individuals. EyeG2P enabled a significant increase in precision in comparison to routine testing strategies (*p*<0.001), with an increased precision in variant analysis of 35% per individual, on average.

**Conclusion:** Automated filtering of genomic variants through EyeG2P can increase the efficiency of diagnostic testing for individuals with a broad range of inherited ophthalmic disorders.

## Introduction

Inherited ophthalmic disorders are a major cause of blindness in children and working age adults.^1,2^ Obtaining a genetic diagnosis in affected individuals can inform management, and clinical genomic testing is increasingly being used as a frontline diagnostic tool for these disorders.^3-8^ Notably, the more widespread availability of gene-directed interventions including gene therapy and preimplantation genetic testing has increased both the value and risk of genomic testing.^9-14^ This places substantial demands on the delivery of testing in a timely and accurate manner.

Here we describe and evaluate the diagnostic utility of EyeG2P, a publically available resource for the analysis of genomic variants identified in genes known as a cause of inherited ophthalmic conditions. We curated disease-causing genes through robust and transparent standards and assessed the sensitivity and precision of EyeG2P in a cohort of individuals receiving diagnostic testing for a range of ophthalmic conditions. EyeG2P uses logical filtering of identified genomic variants in line with their predicted molecular consequence, population frequency and prior knowledge of disease mechanisms and inheritance patterns. Overall we found that utilizing EyeG2P as a first-tier analysis strategy reduces the number of variants requiring analysis by clinically accredited scientists and increases the precision and efficiency of diagnostic testing for ophthalmic disorders.

## Methods

### Curation of known disease genes

The G2P web portal (https://www.ebi.ac.uk/gene2phenotype/)^15^ was used to develop and curate the ophthalmic disorders panel. New entries were initiated by selection of a relevant gene symbol from the list of preloaded genes (with their associated Ensembl identifiers). For each entry, a gene or locus was linked, via a disease mechanism, to a disease. These connections were made after inspecting MEDLINE (through the PubMed interface); search terms included the gene name (HGNC) and the disease name (as a minimum). A disease mechanism was defined as both an allelic requirement (mode of inheritance, for example biallelic or monoallelic) and a mutation consequence (mode of pathogenicity, for example loss-of-function). A confidence attribute—confirmed, probable or possible—was also assigned to indicate how likely it is that the gene is implicated in the cause of disease; the rules used to assign confidence, allelic requirement and mutation consequence to entries are defined and available.^15^ Each locus-genotype-mechanism-disease-evidence link was further characterized by assigning to it a set of phenotype terms (*i*.*e*. clinical signs and symptoms) from the Human Phenotype Ontology (HPO).^16^ The details of the edits and the identifiers of the relevant publications (that provide evidence for that specific gene-disease thread) were stored and are available through the G2P web portal.

### Sequencing and variant identification

All genomic sequencing datasets were generated in a tertiary healthcare setting (North West Genomic Laboratory Hub, Manchester, UK; ISO 15189:2012; UKAS Medical reference 9865). Individuals provided written consent for genomic analysis and all investigations were conducted in accordance to the tenets of the Declaration of Helsinki. All data collected is part of routine clinical care. Analyses to improve genomic diagnostic services for individuals with inherited ophthalmic conditions, as reported in this study, have been approved by the North West Research Ethics Committee (11/NW/0421 and 15/YH/0365). No individual patient data is reported in this manuscript.

Routine diagnostic gene panel testing was performed as previously described.^3,5,17^ Briefly, DNA samples were processed using Agilent SureSelect (Agilent Technologies, Santa, Clara, CA) target enrichment kits designed to capture selected intronic regions and all protein-coding exons +/-50 base pairs of flanking intronic sequences of selected panels of known disease-genes. The decision on which panel to use was made by the referring clinician (either a consultant ophthalmologist or a consultant clinical geneticist with an interest in ophthalmic genetics).

Sequencing was performed using Illumina HiSeq and NextSeq platforms. Raw sequencing reads were aligned to the GRCh37 reference genome using BWA-mem,^18^ with single nucleotide variants (SNVs) and indels identified using GATK.^19^ Larger and more complex indels were identified using Pindel, and copy number variants (CNVs) were identified using DeCON.^20^ Variants were filtered using quality and read depth thresholds as well as inhouse allele frequencies. The zygosity of CNVs were estimated based on their relative read depths. Regions that are highly polymorphic and/or difficult to survey through short-read high-throughput techniques are masked from initial analysis, specifically *RP1L1* exon 4, *USH1C* exon 18 and *RPGR*orf15.

### Variant analysis

#### Routine diagnostics

Routine genomic analysis was performed utilizing the Congenica platform. This process involves filtering variants based on gene/location depending on the gene panel applied, population frequency and predicted molecular consequence. A complete list of presets for variant filtering are available in Supplementary Tables 1&2. After pre-filtering, variants were analysed by clinically accredited scientists and variants classified in accordance with the 2015 American College of Medical Genetics and Genomics (ACMG) best practice guidelines.^21,22^

#### EyeG2P

Merged VCF files containg SNVs, indels and CNVs were annotated using the G2P plugin for Ensembl Variant Effect Predictor.^15,23^ This plugin requires an input file which lists genes of interest and their allelic requirements; we utilized the EyeG2P dataset and an allele frequency cutoff of 0.001 for variants in monoallelic genes and 0.05 for variants in biallelic genes. An additional list was used as input including all ClinVar pathogenic or likely pathogenic variants, all variants predicted to have a significant impact on splicing by SpliceAI,^24^ and a selection hypomorphic alleles that are known to be pathogenic but exceed the variant frequency thresholds specified.

#### Comparisons between routine diagnostic analysis and EyeG2P

Results from EyeG2P were restrospectively compared to clinically reported variants identified from routine diagnostic analysis in 1234 individuals with genomic opthalmic conditions. All these study participants had a confirmed (or a provisional) molecular diagnosis and carried pathogenic (ACMG class 5), likely pathogenic (ACMG class 4) or variants of uncertain significance (ACMG class 3); these changes were identified in a disease-causing state and were deemed to fully account for the patient’s phenotype at the time of routine diagnostic analysis. Prospectively collected data from 83 individuals were also used for comparison. We assessed the sensitivity and the precision of EyeG2P in comparison to results from routine diagnostic analysis. All statistical analyses were performed in R and graphics created in R and BioRender.

## Results

### Curation of the literature identified 667 genes for inclusion in EyeG2P

Between April 2017 and June 2020, we interogated the biomedical literature for genes associated with highly penetrant genetic ophthalmic disorders. We identified 667 unique disease-implicated genes, encompassing 564 MIM disease terms. We weighted the evidence of gene-disease associations from 1624 scientific publications, identifying 559 as ‘confirmed’, 135 as ‘probable’ and 108 as ‘possible’. Within the 559 confirmed gene-disease pairs, the associated inheritance patterns were autosomal dominant in 155, autosomal recessive in 341, X-linked in 31, and other patterns (including both autosomal dominant and recessive) in 32 instances. A high-level assessment of the disease mechanism was performed for each gene-disease combination; among the confirmed pairs, 405 disorders were deemed to be associated with a loss of function mechanism (‘loss of function disorders’), 19 were due to a dominant negative mechanism (‘dominant negative disorders’), and 62 were caused exclusively by missense or inframe insertions-deletion (indel) variants.

The predominantly involved compartment of the eye was determined for the studied gene-disease pairs; dysfunction of the following was identified: retina (*n*=303), lens (*n*=120), cornea (*n*=65), vitreous (*n*=22) and optic nerve (*n*=39). Notably, 245 of the curated pairs were associated with multi-systemic disorders; skeletal (*n*=97), skin (*n*=49), ear (*n*=63), kidney (*n*=48) and metabolism (*n*=39) were the most frequently associated extraocular manifestations.

### EyeG2P is highly sensitive for the detection of disease-causing variants

We assessed the capability of EyeG2P to identify molecular diagnoses in 1234 individuals who had previously undergone diagnostic genetic testing at the North West Genomic Laboratory Hub (Manchester, UK).

EyeG2P was able to prioritize the causal variants in 1228 of the 1234 study participants (99.5%). The 1267 variants prioritized by EyeG2P were identified in 166 distinct genes and had diverse predicted molecular consequences (Figure 1). These variants were detected in 497 individuals with autosomal recessive, 514 individuals with autosomal dominant and 217 individuals with X-linked disorders. The 6 variants missed by EyeG2P included 5’UTR variants and intronic variants not prioritized by SpliceAI.

**Figure 1.**
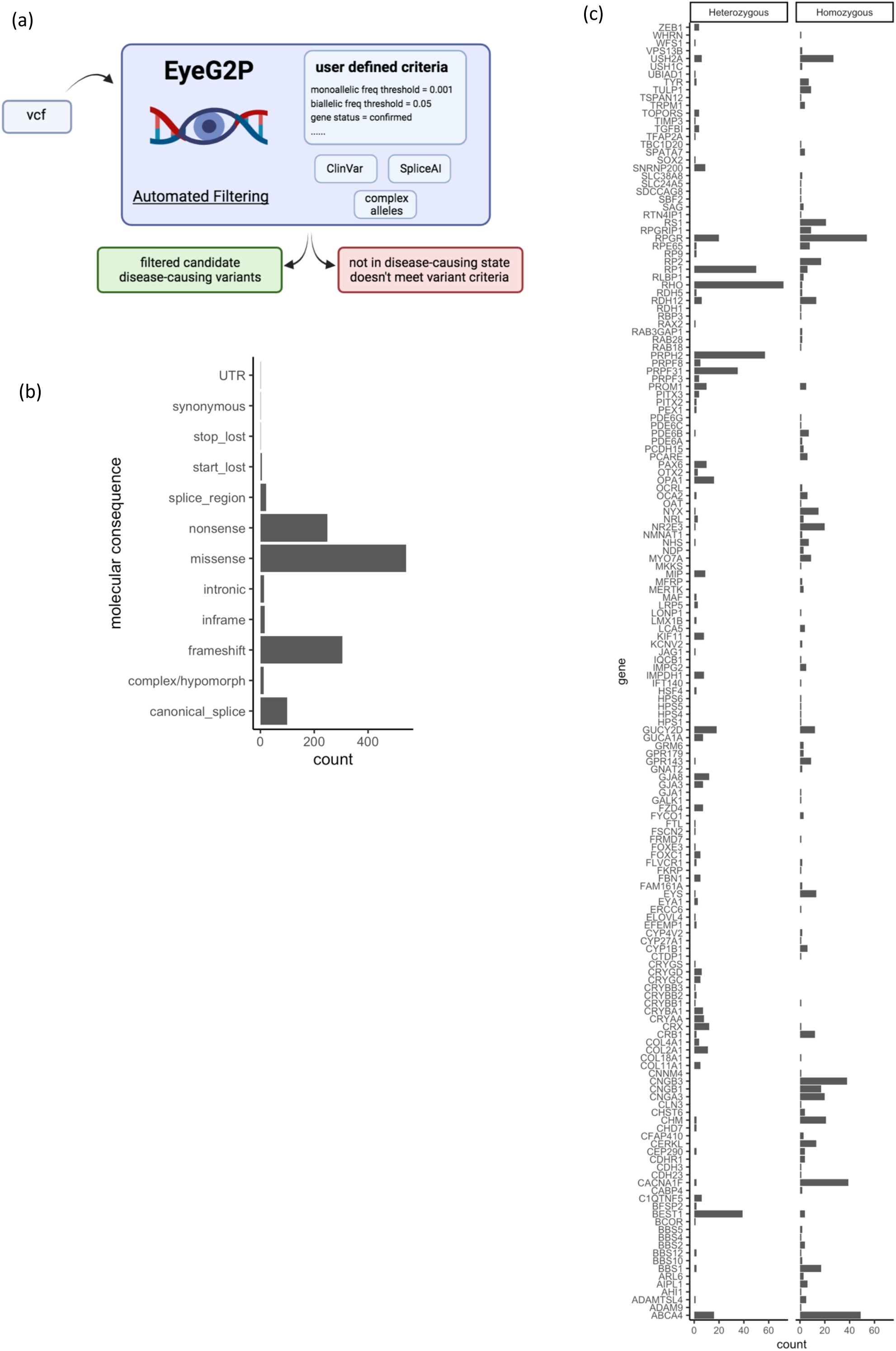
Molecular findings for 1228 individuals with a confirmed molecular diagnosis for inherited Opthalmic disorders. (a) EyeG2P is a plugin for the Ensembl Variant Effector enabling automated variant filtering and selection of variants in a disease-causing state. The specific requirements of variants to be retained can be set by the user and through the inclusion of predefined list of genomic variants, including pathogenic variants in ClinVar, variants predicted to impact splicing, and complex alleles comprised of variants above the defined variant frequency threshold. (b) The predicted molecular consequences of 1267 variants identified as a cause of disease in 1228 individuals, demonstrating the wide range of variant consequences that can be prioritized by EyeG2P. (c) The number of variants identified as a cause of disease in 166 genes by their proven zygosity. Hemizygous variants are included in the *Homozygous* display.

### EyeG2P increases precision over routine diagnostic testing

We performed a prospective and direct head-to-head comparison of EyeG2P to routine diagnostic analysis in an additional test cohort including 83 consecutively ascertained individuals with ophthalmic disease. A confirmed molecular diagnosis was identified for 33/83 cases (40%).

For 31/33 individuals (94%), the confirmed molecular diagnosis was highlighted by EyeG2P; an average reduction of 7.4 variants for analysis was possible in each individual (Figure 2a). Disease-causing variants were identified in 24 distinct genes; 10 cases had an autosomal dominant, 3 an X-linked and 18 an autosomal recessive disorder. The genomic variants underpinning these diagnoses included 3 CNVs (exonic deletions), 5 indels and 35 SNVs (Figure 2b). In the remaining 52/83 individuals, we identified 2 cases with a confirmed genetic diagnosis as a result of variants in the orf15 region of the *RPGR* gene; *RPGR*orf15 was excluded from EyeG2P analysis due to difficulties of short-read sequencing approaches to accurately identify variants in this region. In the remaining 50/52 (96.2%) cases without a molecular diagnosis following EyeG2P analysis, we were unable to identify a confirmed diagnosis through routine genetic testing approaches, with an increased analysis burden of 6.1 variants, on average, per individual (Figure 2a). In 10/52 cases without a confirmed diagnosis, variants of uncertain significance were identified in a disease-causing state through EyeG2P analysis, and no additional pathogenic variants were detected after routine analysis.

**Figure 2.**
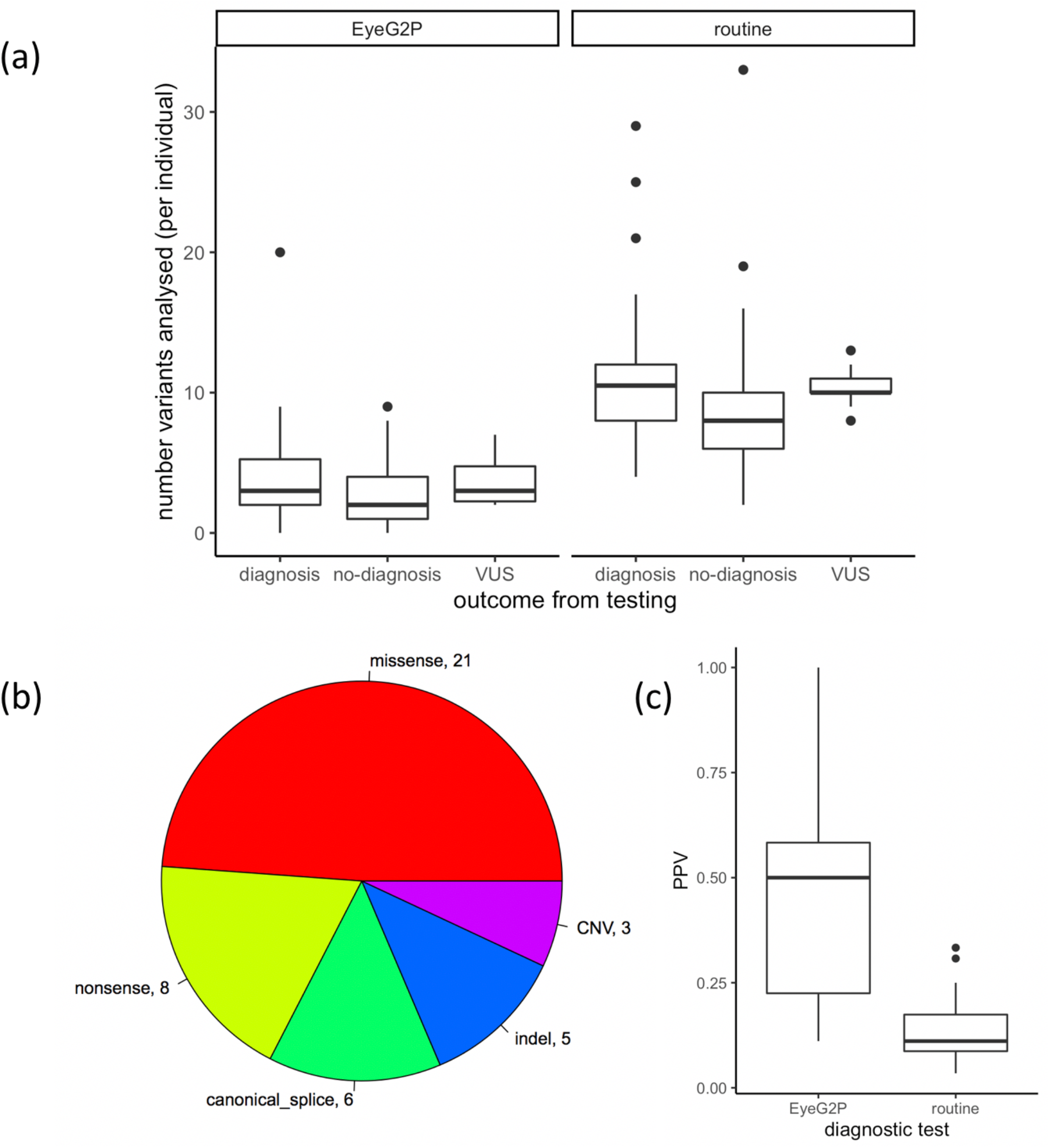
Direct comparison between EyeG2P and routine testing approaches identifies increased precision and efficiency of EyeG2P as a first-tier analysis approach. (a) the number of variants requiring analysis by clinical scientists for 83 individuals receiving genetic testing for inherited Ophthalmic disorders. (b) summary of the molecular consequences of disease-causing variants identified to underpin confirmed molecular diagnoses in 31 individuals. (c) the precision (PPV, positive predictive value) of different testing approaches for 31 individuals receiving a diagnosis through both approaches.

We calculated the precision (positive predictive value, PPV) of genetic testing through EyeG2P and routine diagnostic genetic testing procedures for the 31 individuals who received a molecular diagnosis through both approaches. We observed a significant increase in precision in EyeG2P testing compared to default testing strategies (Paired Wilcoxon Rank Sum Test, *p*<0.001), with an average increase in precision of 35% (Figure 2c).

## Discussion

Characterising the genomic basis of inherited ophthalmic conditions has been shown to inform the management of individuals with these conditions.^3,9^ The expansion from single gene based testing methodologies to the routine use of large gene panels, exome and genome sequencing approaches requires that robust and accurate informatics filtering strategies are applied to the generated datasets. Moreover, the continual identification of new disease-associated genes and disease mechanisms requires sequencing and analytical approaches that are dynamic and can evolve with expanding knowledge. Here, we described EyeG2P, a filtering approach available as a plugin for the Ensembl Variant Effect Predictor.^15^ EyeG2P can be applied to any genomic variant dataset in VCF format, including targeted gene panels, exome and genome datasets. We show that EyeG2P increases the precision and efficiency of genomic testing for inherited ophthalmic conditions over routine approaches for variant analysis, at little cost to overall diagnostic rates.

The genetic basis of ophthalmic conditions such as congenital cataract, inherited retinal disorders and optic neuropathies is diverse and includes genes encoded on autosomal, sex and mitochondrial chromosomes. Following curation of over 1000 biomedical publications we identified 667 relevant genes and determined the associated modes of inheritance, mechanisms of disease causation and phenotypic features. We have released these data as a freely available resource that can be dynamically filtered and revised to best aid the users requirements. For example, the recent elucidation of *DYNC2H1* as a cause of inherited ophthalmic conditions was not captured in our initial curation process but can be subsequently included in EyeG2P analysis through addition of a single data line to the released EyeG2P datafile.^25^

Our ability to detect disease-causing genomic variants from high-throughput sequencing datasets has expanded in recent years to include complex structural variants,^26,27^ exonic deletions and duplications,^28,29^ deeply intronic variants causing aberrant splicing,^30-33^ variants in regulatory regions^34-36^ and complex alleles comprised of combinations of genomic variants common in the general population.^37,38^ Here we found that, in addition to characterizing novel exonic variants, EyeG2P is capable of prioritizing these diverse types of disease-causing variation, achieving 99.5% sensitivity in comparison to routine analytical approaches (Figure 1). As our knowledge of the specific genomic variants causing these conditions expands, it is possible for the user to adjust the analysis settings of EyeG2P to meet these requirements and/or provide a list of specific variants for inclusion. Such approaches can be applied at scale and are useful, for example, to identify novel deeply intronic variants predicted to impact splicing that are in a disease-causing state with other intronic or exonic variants.

In conclusion, we demonstrate that EyeG2P can be effectively integrated with clinical diagnostic testing for inherited ophthalmic conditions to increase the efficiency of variant analysis. We show that EyeG2P reduces the variant analysis workload for clinical scientists and increases the precision of diagnostic testing. Moreover, EyeG2P can identify diverse genomic variants across the spectrum of genetically and clinically heterogeneous ophthalmic genetic conditions. We propose the application of EyeG2P as a first-tier analysis strategy for the diagnosis of inherited opthalmic conditions from high-throughput genomic datasets.

## Data Availability

The EyeG2P dataset is available from https://www.ebi.ac.uk/gene2phenotype/downloads.

## Data Availability

The EyeG2P dataset is available from https://www.ebi.ac.uk/gene2phenotype/downloads.

## Acknowledgements

We thank the families, clinicians, genetic counsellors, laboratory staff, clinical scientists and bioinformaticians involved in the routine diagnostic analysis for individuals included in this study.

Ensembl receives majority funding from Wellcome (grant numbers WT095908, WT098051, WT108749/Z/15/Z). We acknowledge funding from the Wellcome Trust Transforming Genomic Medicine Initiative (WT200990/Z/16/Z), the European Molecular Biology Laboratory and the Manchester NIHR Biomedical Research Centre (IS-BRC-1215-20007). JME is funded by a postdoctoral research fellowship from the Health Education England Genomics Education Programme.

---

**Supp Table 1:**
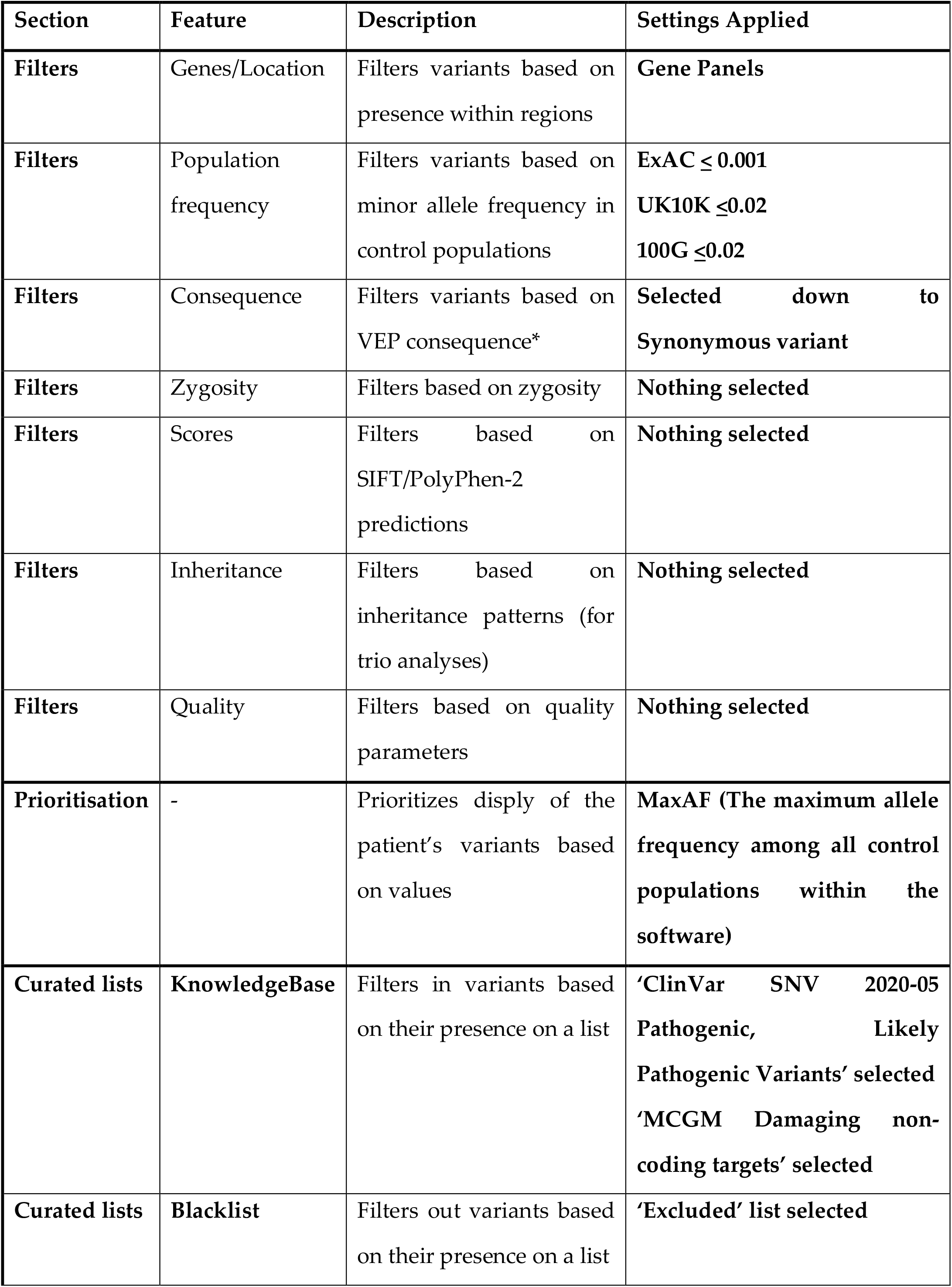
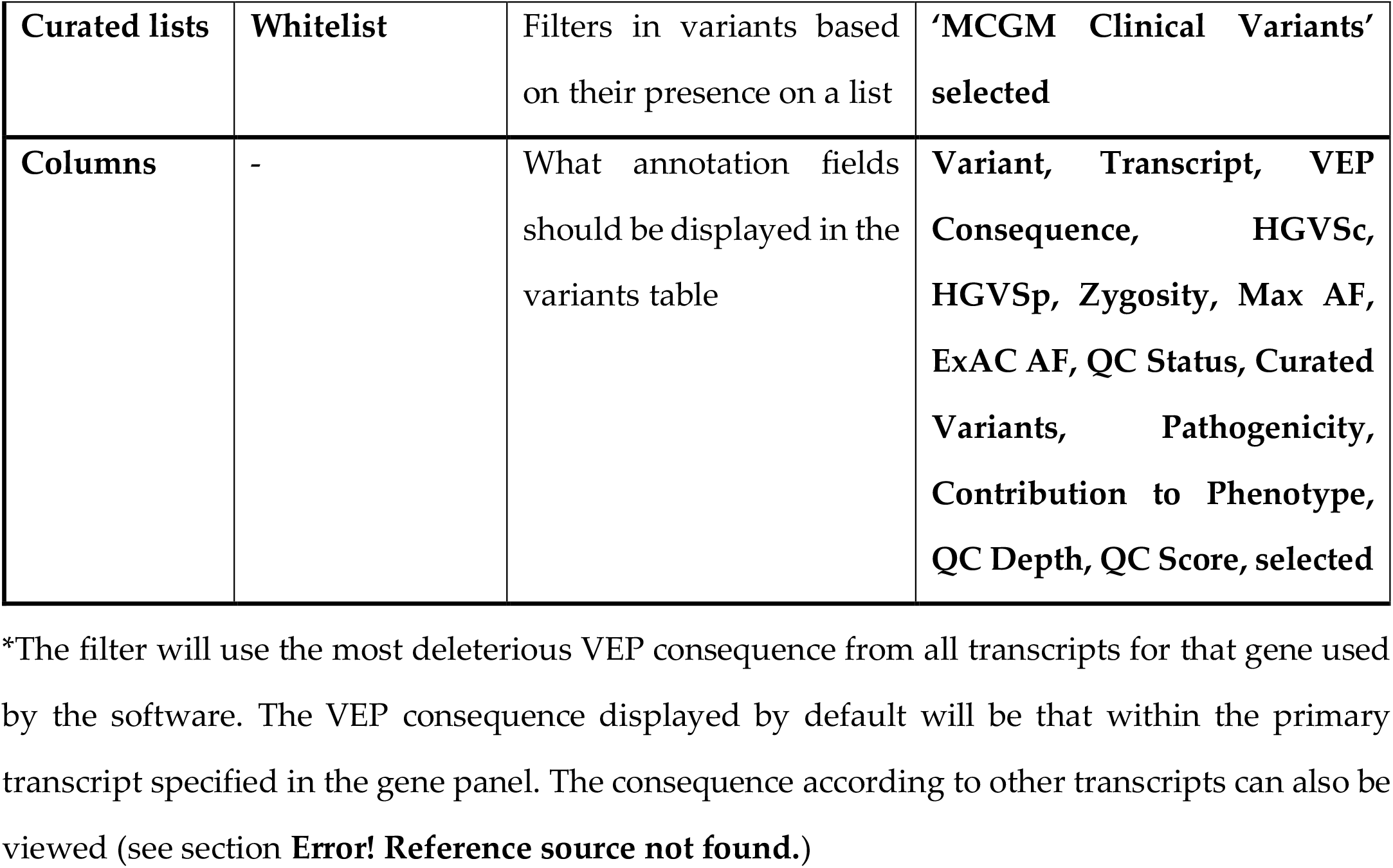
Filter presets for analysis in Congenica platform for coding SNVs

| Section | Feature | Description | Settings Applied |
| --- | --- | --- | --- |
| **Filters** | Genes/Location | Filters variants based on presence within regions | **Gene Panels** |
| **Filters** | Population frequency | Filters variants based on minor allele frequency in control populations | **ExAC $\leq 0.001$**<br>**UK10K $\leq 0.02$**<br>**100G $\leq 0.02$** |
| **Filters** | Consequence | Filters variants based on VEP consequence* | **Selected down to**<br>**Synonymous variant** |
| **Filters** | Zygosity | Filters based on zygosity | **Nothing selected** |
| **Filters** | Scores | Filters based on SIFT/PolyPhen-2 predictions | **Nothing selected** |
| **Filters** | Inheritance | Filters based on inheritance patterns (for trio analyses) | **Nothing selected** |
| **Filters** | Quality | Filters based on quality parameters | **Nothing selected** |
| **Prioritisation** | - | Prioritizes display of the patient's variants based on values | **MaxAF (The maximum allele frequency among all control populations within the software)** |
| **Curated lists** | **KnowledgeBase** | Filters in variants based on their presence on a list | **'ClinVar SNV 2020-05 Pathogenic, Likely Pathogenic Variants' selected**<br>**'MCGM Damaging non-coding targets' selected** |
| **Curated lists** | **Blacklist** | Filters out variants based on their presence on a list | **'Excluded' list selected** |
| **Curated lists** | **Whitelist** | Filters in variants based on their presence on a list | **'MCGM Clinical Variants' selected** |
| **Columns** | - | What annotation fields should be displayed in the variants table | **Variant, Transcript, VEP Consequence, HGVS<sub>c</sub>, HGVS<sub>p</sub>, Zygosity, Max AF, ExAC AF, QC Status, Curated Variants, Pathogenicity, Contribution to Phenotype, QC Depth, QC Score, selected** |

\*The filter will use the most deleterious VEP consequence from all transcripts for that gene used by the software. The VEP consequence displayed by default will be that within the primary transcript specified in the gene panel. The consequence according to other transcripts can also be viewed (see section **Error! Reference source not found.**)

Where projects require different filters applying as standard, this should be specified in the specific service profile.

**Supp Table 1:**
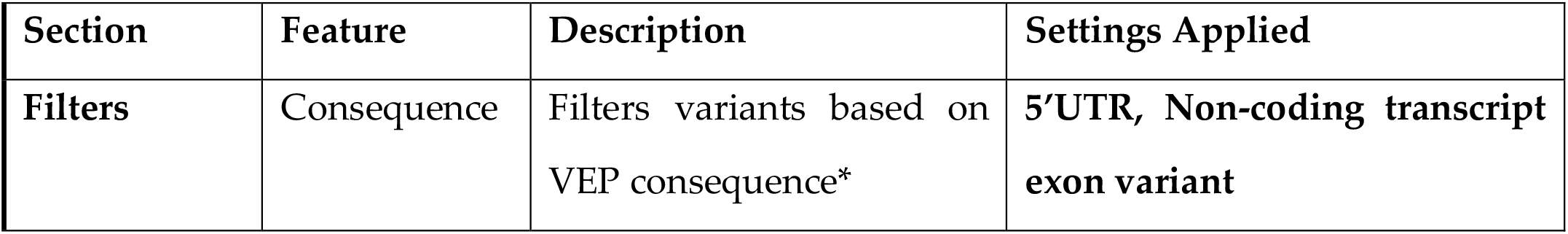
Filter presets for analysis in Congenica for non-coding SNVs

| **Section** | **Feature** | **Description** | **Settings Applied** |
| --- | --- | --- | --- |
| **Filters** | Consequence | Filters variants based on VEP consequence* | **5'UTR, Non-coding transcript exon variant** |

## Notes

### Competing Interest Statement

The authors have declared no competing interest.

